# A novel *Sporothrix brasiliensis* genomic variant in Midwestern Brazil: evidence for an older and wider sporotrichosis outbreak

**DOI:** 10.1101/2020.07.09.20142125

**Authors:** João Eudes Filho, Isabele Barbieri Santos, Carmélia Matos Santiago Reis, José Salvatori Patané, Verenice Paredes, João Paulo Romualdo, Sabrina Santos Costa Poggianni, Talita de Cássia Borges Castro, Oscar Mauricio Gomez, Sandro Antonio Pereira, Edvar Yuri Pacheco Schubach, Kamila Peres Gomes, Heidi Mavengere, Lucas Gomes de Brito Alves, Joaquim Lucas, Hugo Costa Paes, Patricia Albuquerque, Laurício Monteiro Cruz, Juan G. McEwen, Jason E. Stajich, Rodrigo Almeida-Paes, Rosely Maria Zancopé-Oliveira, Daniel R. Matute, Bridget Barker, Maria Sueli Soares Felipe, Marcus de Melo Teixeira, André Moraes Nicola

**Author notes:** **Corresponding authors:** Name: André Moraes Nicola, Address: Campus Universitário Darcy Ribeiro, Núcleo de Medicina Tropical, sala 40. Brasília- DF, Brazil. 70910-900., Name: Marcus de Melo Teixeira, Address: Campus Universitário Darcy Ribeiro, Núcleo de Medicina Tropical, sala 40. Brasília- DF, Brazil. 70910-900. These authors share senior authorship (Marcus de Melo Teixeira, André Moraes Nicola).

## Abstract

Sporotrichosis is a subcutaneous infection caused by fungi from the genus *Sporothrix*. The disease is transmitted by inoculation of infective particles found in plant-contaminated material or diseased animals, characterizing the classic sapronotic and emerging zoonotic transmission, respectively. Since 1998, Brazil has experienced a zoonotic sporotrichosis epidemic due to *S. brasiliensis*, centered in the state of Rio de Janeiro. Our observation of feline sporotrichosis cases in Brasília (Midwestern Brazil), around 900 km away from Rio de Janeiro, led us to question whether the epidemic caused by *S. brasiliensis* has spread from the epicenter in Rio de Janeiro, emerged independently in the two locations, or whether the disease has been present and unrecognized in Midwestern Brazil. A retrospective analysis of 91 human and 4 animal cases from Brasília, ranging from 1993 to 2018, suggests the occurrence of both sapronotic and zoonotic transmission. Molecular typing identified *S. schenckii* as the agent in two animals and all seven human patients from which we were able to recover clinical isolates. However, in two animals, the disease was caused by *S. brasiliensis*. Whole-genome sequence typing of seven *S. brasiliensis* strains suggests that isolates from Brasília are genetically distinct from those obtained at the epicenter of the outbreak in Rio de Janeiro, both in phylogenomic and population genomic analyses. The two *S. brasiliensis* populations seem to have separated 2.24 - 3.09 million years ago, indicating independent outbreaks or that the zoonotic *S. brasiliensis* outbreak might have started earlier and be spread wider in South America than previously recognized.

## Introduction

The emergence of fungal pathogens worldwide is an important and frequently neglected public health issue [1]. One of the most frequent genera of pathogenic fungi is *Sporothrix* (Sordariomycetes, Ascomycota), which causes the subcutaneous granulomatous disease called sporotrichosis. [2,3]. The etiologic agent of the disease, *S. schenckii* sensu lato, was recently described to harbor three cryptic species: *S. schenckii* sensu stricto, *S. globosa*, and the emerging species *S. brasiliensis* [4-6]. The three species are saprophytes and produce sympodial and sessile conidia that switch their morphology to the pathogenic yeast phase after a transcutaneous injury [7]. The three species differ in their virulence. *S. brasiliensis* is more virulent in animal models than *S. schenckii* and *S. globosa*, and causes localized to disseminated sporotrichosis that can be fatal to both humans and cats [8]. Lower antifungal susceptibility and refractory sporotrichosis occurs in some Brazilian cases related to the zoonotic epidemics [9,10]. Previous population genetics surveys revealed that *S. brasiliensis* is largely clonal and the little variation in the species is associated to particular geographic areas [6]. On the other hand, its sister taxon *S. schenckii* shows higher rates of recombination and little to no population structure among Brazilian strains [6].

Currently, sporotrichosis occurs endemically globally but it is especially prevalent in Latin America, South Africa, Madagascar, China, India, and Japan [3]. In addition to endemic disease, multiple sporotrichosis outbreaks have been reported in the last century [2,11,12]. Before the 1990s, sporotrichosis was considered a disease of low-to-moderate endemicity in Brazil. In the past two decades, though, it has become an urgent health problem [7,13]. In 1997, three individuals from the same family got infected by *Sporothrix* sp. after successive feline zoonotic transmission events in Rio de Janeiro, Brazil [14]. Since 1998, over 5,100 cats have been diagnosed with sporotrichosis in the state of Rio de Janeiro; >5,000 human cases were reported in 2015, suggesting high prevalence of the disease in the state [15]. Rio de Janeiro is currently considered one of the three (along with China and South Africa) sporotrichosis hyperendemic regions [14,16,17], with a crucial difference in incidence of feline sporotrichosis and zoonotic transmission from cats to humans [6,18].

Infections with *S. brasiliensis* have recently been described in Argentina and Paraguay, the first reports of this species outside Brazil [19,20]. Sporotrichosis caused by *S. brasiliensis* has recently been described in Northeast Brazil, also mostly due to zoonotic transmission [21-24]. The recent emergence of *S. brasiliensis-*associated sporotrichosis in places with few or no cases reported in the early 2000s raises the question of whether the outbreak has recently spread from Rio de Janeiro or zoonotic sporotrichosis cases had been happening there, unrecognized. This question is very important in determining public health policy do deal with emergent sporotrichosis. In this work, prompted by our observation in 2015 of a case of sporotrichosis in a cat whose owner also had a lesion that was suggestive of the disease, we address this question.

To look for evidence of human zoonotic infections, we documented additional animal cases and retrospectively studied a series of sporotrichosis cases in the University Hospital of Brasília. We also sequenced the genomes of seven *Sporothrix* spp. strains and compared them to the reference *S. brasiliensis* 5110 genome [25]. We report for the first time the occurrence of *S. brasiliensis* infections in animals at the capital of Brazil, Brasília, located in the Midwestern portion of the country. We also found a surprising genomic diversity in the *S. brasiliensis* strains in Brasília, which suggests that independent outbreaks caused by *S. brasiliensis* occur and might have started earlier and be spread wider than previously recognized.

## Material and Methods

### Human cases

We carried out a cross-sectional study of a series of sporotrichosis cases in the University Hospital of Brasília, from 1993 to 2018. This study was previously approved by the Ethics Committee of the Faculty of Medicine, University of Brasília (protocol CAAE: 873718.0.00005558). The inclusion criterion was diagnosis of sporotrichosis by culture of *Sporothrix* sp. in the Mycology Laboratory of the University Hospital of Brasília. Diagnoses were performed by growing the fungus from biological samples (skin lesions, sputum, broncho-alveolar lavage, and cerebrospinal fluid) on solid culture media (i.e. Sabouraud and Mycosel) followed by microscopic characterization of colonies. The following data were anonymously retrieved from patient medical records: clinical form of sporotrichosis (cutaneous, mucosal and extracutaneous forms), address, year of diagnosis, age, sex, gender, race, occupation, treatment, and disease outcome.

### Animal cases

Three cats and one dog with suspected sporotrichosis from the Veterinary Hospital of the University of Brasília, the Directorate for Environmental Surveillance (zoonosis division) of the Federal District Health Department and a private veterinary clinic were referred to the Mycology Laboratory of the University Hospital of Brasília for diagnosis between 2015 and 2018. We performed direct microscopic observation of patient samples and cultured skin lesion specimens as above, followed by microscopic identification of the fungus. Relevant clinical and epidemiological data were also collected to describe the veterinary cases. Animal observations and tests were previously approved by the University of Brasília Ethics Committee on the Use of Animals (CEUA-UnB), protocol: 66716/2016.

### Morphological studies

Seven isolates from humans and four isolates from animal cases were used for morphological and molecular characterization. Pure cultures were kept on Sabouraud dextrose agar at room temperature for the development and identification of the mycelial phase. The slide culture method was used to characterize microscopic filamentous features of the *Sporothrix* spp. isolates. Mycelia fragments were inoculated into 1 x 1 cm Sabouraud dextrose agar and malt extract agar blocks, which were then mounted on a slide, covered with a coverslip and incubated for 14-21 days at room temperature. To induce the yeast phase, the mycelial fragments were inoculated into Brain-Heart Infusion broth and incubated for 7 days at 37 °C and 150 rpm agitation. Both mycelial and yeast preparations were stained with lactophenol blue and visualized with a Zeiss AxioObserver Z1 microscope equipped with a 40X objective. Images were collected using the Zeiss ZEN software.

### Molecular typing and identification

We extracted genomic DNA using the PureLink Genomic DNA Mini Kit (ThermoFisher). We started with 500 mg of mycelial tissue as input material. We used glass beads protocol with a Precellys 24 instrument (Bertin). We assess the DNA integrity by running 5 μL aliquot of the extraction on a in a 0.8% agar agarose-gel electrophoresis stained with 0.5 µg/ml ethidium bromide. To estimate the DNA yield were we used a NanoDrop 1000c Instrument (ThermoFisher). We then amplified a segment of the calmodulin gene with PCR and Sanger sequenced in an ABI 3130xl Instrument (Applied Biosystems). PCR setting and cycling conditions were as previously described [6]. Initial nucleotide BLAST analysis (BLASTn) was performed for each amplicon in order to retrieve the best hit results for each strain on the NCBI database. All extractions (N=15) showed *Sporothrix* spp. as the closest BLAST match.

### Whole genome sequencing

We selected six *S. brasiliensis* (A001, A005, s15677, s28606, s34180 and s48605) and one *S. schenckii* (A003) isolates for Illumina short read whole-genome sequencing. The two *S. brasiliensis* strains, A001 and A005, were retrieved from the collection at the Mycology Laboratory of the University Hospital of Brasília and labelled as SbFD (*S. brasiliensis*, Federal District). Isolates s15677, s28606, s34180 and s48605 were from the Collection of Pathogenic Fungi at the Oswaldo Cruz Foundation, were obtained in the state of Rio de Janeiro and were designated as SbRJ. From each isolate, we extracted 1 µg of DNA, as described above. Next, we prepared paired-end libraries using the NEBNext® Ultra™ DNA Library Prep Kit for Illumina. The libraries were quantified using the NEBNext® Library Quant Kit for Illumina® (New England Biolabs). We pooled equimolar amounts of each library together and sequenced them using in an Illumina NextSeq 550 Instrument using the NextSeq 500/550 High Output Kit v2 chemistry. The run was performed in a high output, 2 × 150 bp mode. We used Trimmommatic [26] to remove adapters and filter low-quality bases.

### Public data

We used three additional sets of Illumina paired-end reads from *S. schenckii* isolates from Colombia (EM7 and MS1 - [27]) and Puerto Rico (ATCC 58251 - [28]). To root phylogenetic trees (see below), we retrieved additional genomic assemblies from other *Sporothrix* species from the NCBI genome browser (Supplementary Table 1).

**Table 1.**
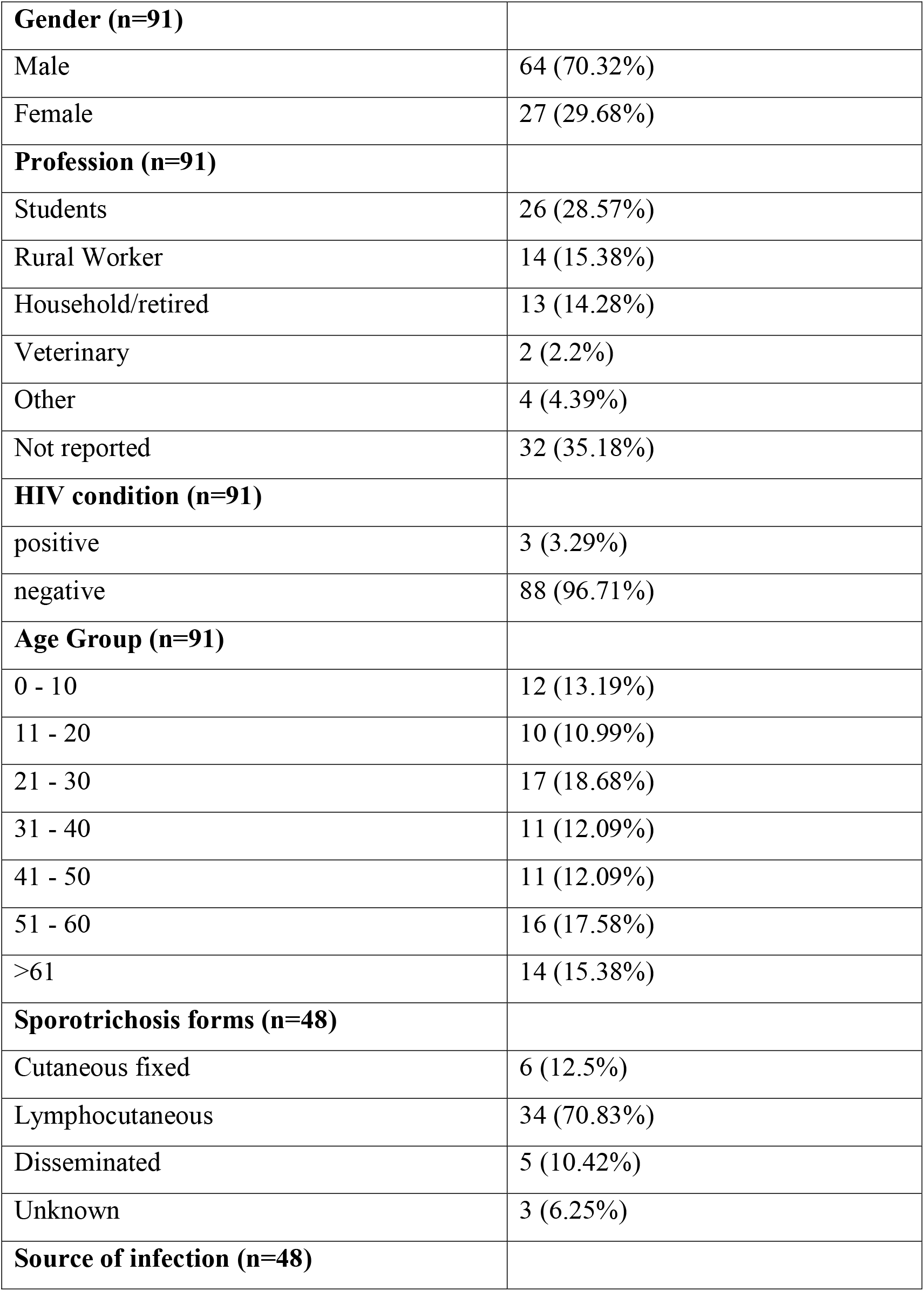
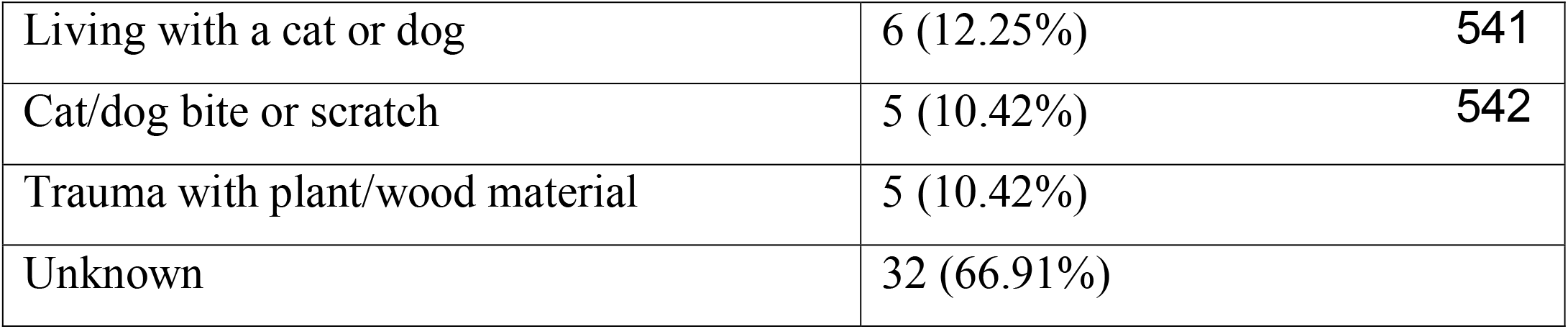
Epidemiological characteristics of sporotrichosis in Brasília, Brazil.

|  |  |
| --- | --- |
| **Gender (n=91)** |  |
| Male | 64 (70.32%) |
| Female | 27 (29.68%) |
| **Profession (n=91)** |  |
| Students | 26 (28.57%) |
| Rural Worker | 14 (15.38%) |
| Household/retired | 13 (14.28%) |
| Veterinary | 2 (2.2%) |
| Other | 4 (4.39%) |
| Not reported | 32 (35.18%) |
| **HIV condition (n=91)** |  |
| positive | 3 (3.29%) |
| negative | 88 (96.71%) |
| **Age Group (n=91)** |  |
| 0 - 10 | 12 (13.19%) |
| 11 - 20 | 10 (10.99%) |
| 21 - 30 | 17 (18.68%) |
| 31 - 40 | 11 (12.09%) |
| 41 - 50 | 11 (12.09%) |
| 51 - 60 | 16 (17.58%) |
| >61 | 14 (15.38%) |
| **Sporotrichosis forms (n=48)** |  |
| Cutaneous fixed | 6 (12.5%) |
| Lymphocutaneous | 34 (70.83%) |
| Disseminated | 5 (10.42%) |
| Unknown | 3 (6.25%) |
| **Source of infection (n=48)** |  |

|  |  |  |
| --- | --- | --- |
| Living with a cat or dog | 6 (12.25%) | 541 |
| Cat/dog bite or scratch | 5 (10.42%) | 542 |
| Trauma with plant/wood material | 5 (10.42%) |  |
| Unknown | 32 (66.91%) |  |

### Read mapping and variant calling

We mapped the newly-sequenced strains and the publicly available data to the *S. brasiliensis* 5110 reference genome [29] using bwamem. To identify Mismatch intervals and indels, we used GATK v3.3-0RealignerTargetCreator and IndelRealigner [30,31]. To identify polymorphic sites (single nucleotide polymorphisms, SNP), we used the GATK UnifiedGenotyper module using the parameter het = 0.01 to account for a haploid organism. We used the following filters to obtain SNPs calls: QD = 2.0 ‖ FS_filter = 60.0 ‖ MQ_filter = 30.0 ‖ MQ_Rank_Sum_filter = -12.5 ‖ Read_Pos_Rank_Sum_filter = -8 [32]. We excluded polymorphic sites with less than 10X coverage, with less than 90% variant allele calls, or that were identified by Nucmer [33] as located in duplicated regions in the *S. brasiliensis* reference genome. We identified 115,488 SNPs across five *Sporothrix* species (16 isolates).

### Phylogenetic tree and whole-genome alignments

We used the WGS dataset described above to build a phylogenetic tree of the 16 *Sporothrix* spp. isolates. We used IQTREE [34] to build a Maximum Likelihood (ML) tree with the –m TEST function to determine the best nucleotide substitution model. To assess the strength of the support for each clade present in the tree, we found the consensus phylogenomic tree of 1,000 ultrafast bootstraps coupled with 1,000 Shimodaira–Hasegawa-like approximate likelihood ratio tests (SH-aLRT) [35,36]. We visualized the consensus topology and branch support with FigTree v1.4.2 – http://tree.bio.ed.ac.uk/software/figtree/. We used the Automatic Assembly For The Fungi (AAFTF) pipeline to assemble the newly-sequenced *S. brasiliensis* genomes, with default parameters (https://github.com/stajichlab/AAFTF). The *S. brasiliensis* assemblies with the highest N50 (Supplementary Table 1) were aligned to the strain 5110 reference genome using the DGENIES online tool via minimap2 algorithm for comparative genomic purposes.

**Figure 1.**
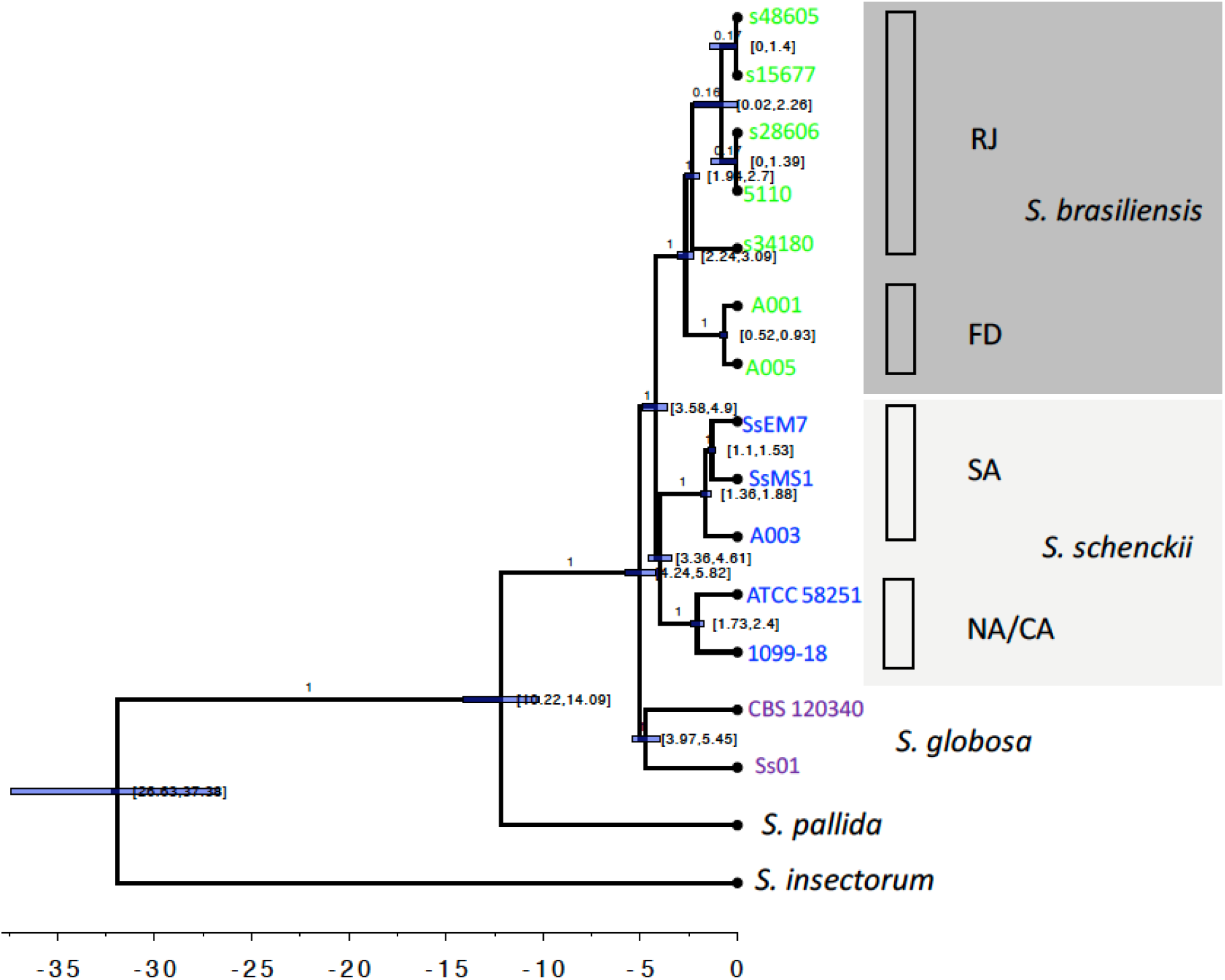
Whole-genome *Sporothrix* genealogy using BEAST analyses. The phylogenomic tree was inferred using Bayesian inference under the GTR model with gamma variation among sites and *S. insectorum* was set as the outgroup. The 90% confidence interval marking the separation between *S. brasiliensis* and *S. schenckii* was set to 3.8 - 4.9 MYA, a rate interval of [0.9E-3, 16.7E-3] based on evolutionary rate estimation based on different groups of fungi and a birth-death tree prior with incomplete sampling were set as priors. The clade distribution, posterior probabilities and divergence times are shown.

#### Timing analysis

Next, we estimated the approximate divergence time of the splits observed in the phylogenomic analysis using Bayesian inference in BEAST v1.10.4 [37]. To avoid overestimation of branch lengths which would tend to inflate molecular rates, a number of invariable sites were input in the .xml files according to their frequency in the species genome’s composition and their base count of each type (A, C, G, and T) in the respective genome. We used a GTR model with gamma variation among sites (using four discrete classes). Priors for the different dating parameters included: a Normal distribution in which a conservative 90% confidence interval was within 3.8 - 4.9 MYA (following [25]) marking the separation between *S. brasiliensis* and *S. schecnkii*; a rate interval of [0.9E-3, 16.7E-3] substitutions/site/branch/MY for the parameter ucld.mean (mean of the uncorrelated lognormal distribution of rates across branches, instead of assuming a more simplistic global clock prior), based on evolutionary rate estimation based on different groups of fungi [38]; and a birth-death tree prior with incomplete sampling [39]. We used two independent Markov Chain Monte Carlo (MCMC) chains each run until convergence; the marginal distributions and traces from the two replicate chains were inspected in Tracer v1.7 [40]. We summarized the two MCMC runs using LogCombiner (BEAST package) after discarding the *burnin* region. Finally, we used Treeannotator (BEAST package) to annotate clade posteriors, divergence times, and rate variation across branches.

#### Population genetic analysis

We estimated the average nucleotide diversity (π) within species or populations and absolute differentiation (*D*_*xy*_) between *S. brasiliensis* and *S. schenckii*, and between the *S. brasiliensis* SbFD and SbRJ using a series of Python scripts available at https://github.com/simonhmartin/genomics_general.

## Results

### Both sapronotic and zoonotic transmissions of sporotrichosis occur in Brasília and surrounding areas

The study population involved 91 human cases ranging from 1 to 82 years of age with sporotrichosis confirmed by isolation of *Sporothrix* spp. in culture at the Mycology Laboratory of the University Hospital of Brasília. All patients were diagnosed either by direct exam or by the microbiological isolation of the pathogen from 1993 to 2018. We reported 64 cases (70%) in males and 27 (30%) females. The years with most cases reported were 1999, 2000, and 2009 with 12 (13%), 7 (8%), and 8 (9%) cases, respectively. Regarding age groups, the 21-30- and 51-60-year ranges have more cases in this retrospective study, corresponding to 17 (18.7%) and 16 (17.6%) cases, respectively (Table 1). Most patients were treated with itraconazole alone or in association with saturated potassium iodide and, in severe cases, fluconazole and/or amphotericin B were used.

We were able to analyze 48 full medical records of the included patients. Among them, 34 were presumably infected in Brasília (70.9%) and six in the neighboring state of Goiás (12.5%). Most patients had the lymphocutaneous form (n=34 – 70.8%), followed by fixed cutaneous form (n=6 – 12.6%). Five extra-cutaneous cases were detected in the central nervous system (n=1 – 2%) and or the upper airway tract (n=4 - 8.3%). The clinical presentation of three patients (6.3%) was inconclusive or not reported. The most frequent profession/occupation was student (n=26 - 29%), followed by farmers and retirees with 15% (n=14) and 14% (n=13), respectively (Table 1). Some risk factors we found were prostate and breast cancer, high blood pressure, diabetes, cirrhosis, tuberculosis, leishmaniosis, hepatitis and other chronic diseases. Chagas disease and paracoccidioidomycosis co-infections were found in two sporotrichosis patients. Three cases of HIV coinfection were also diagnosed. Three cases were fatal, two of which in patients with cancer and one with chronic cirrhosis.

Additionally, a thorough investigation of the clinical records suggested two main forms of *Sporothrix* infections in Brasília: (1) the classical sapronotic form acquired from a direct ninoculation of plant-derived material harboring the fungus, which was more observed in rural and construction workers, and (2) the zoonotic transmission triggered by skin injuries provoked by infected cat or dog bite/scratches. Sapronotic sporotrichosis was reported in 5 patients (10%) after injuries with plant-derived material. Five patients (10%), including two veterinarians, reported cat/dog bites or scratches and six other patients (12.6%) reported living and having frequent contact with dogs or cats with skin lesions. Other patients either did not report any previous injuries or reported a different exposure source (i.e. sporotrichosis acquired after injuries with barbed wire and metal cans).

In parallel, we diagnosed, from 2015 to 2018, 4 animals (3 cats and 1 dog) with sporotrichosis in Brasília. These animals had classical clinical presentations such as multiple suppurative granulomatous cutaneous lesions with a high load of yeast cells, generally with involvement of mucous membranes. Lesions were commonly found at parts such as the head, especially on the nose, as well as on the chest and legs. Two felines were euthanized and the other cat and the dog were treated with itraconazole and potassium iodide. Lastly, the pet owner of the cat in which the A001 strain was recovered, was also diagnosed with cutaneous sporotrichosis. The diagnosis and treatment were made in another hospital in Brasília, so we do not have medical records or a clinical isolate obtained from the owner.

### Sporothrix brasiliensis isolates from Brasília and the epicenter of the zoonotic sporotrichosis epidemics, Rio de Janeiro, are genetically distinct

In order to differentiate between two different hypotheses for *S. brasiliensis* presence in Brasília, that it was recently introduced here from Southeast Brazil or that the Brasília isolates are actually endemic to that region, we performed a number of genetics and genomics analyses. First, we inferred the genealogical relationships between isolates of different species of *Sporothrix* using whole genome sequencing. By rooting our tree with *S. insectorum*, we observed that *S. schenckii* and *S. brasiliensis* are sister species and form a triad along with *S. globosa* (Figure 2). The Most Recent Common Ancestor (tMRCA) of *Sporothrix* emerged between 26.63 and 37.38 Million Years Ago (MYA) (Figure 3). Unlike *S. insectorum* and *S. pallida*, this triad of species cause sporotrichosis in humans and animals and form the “*Sporothrix* pathogenic clade”. The crown age of this pathogenic group (TMRCA) is between 4.24 - 5.28 MYA. The divergence between the pathogenic clade and *S. pallida* occurred 10.22 - 14.09 MYA, suggesting that the pathogenesis syndrome must have arisen in the last 15 MY.

**Figure 2.**
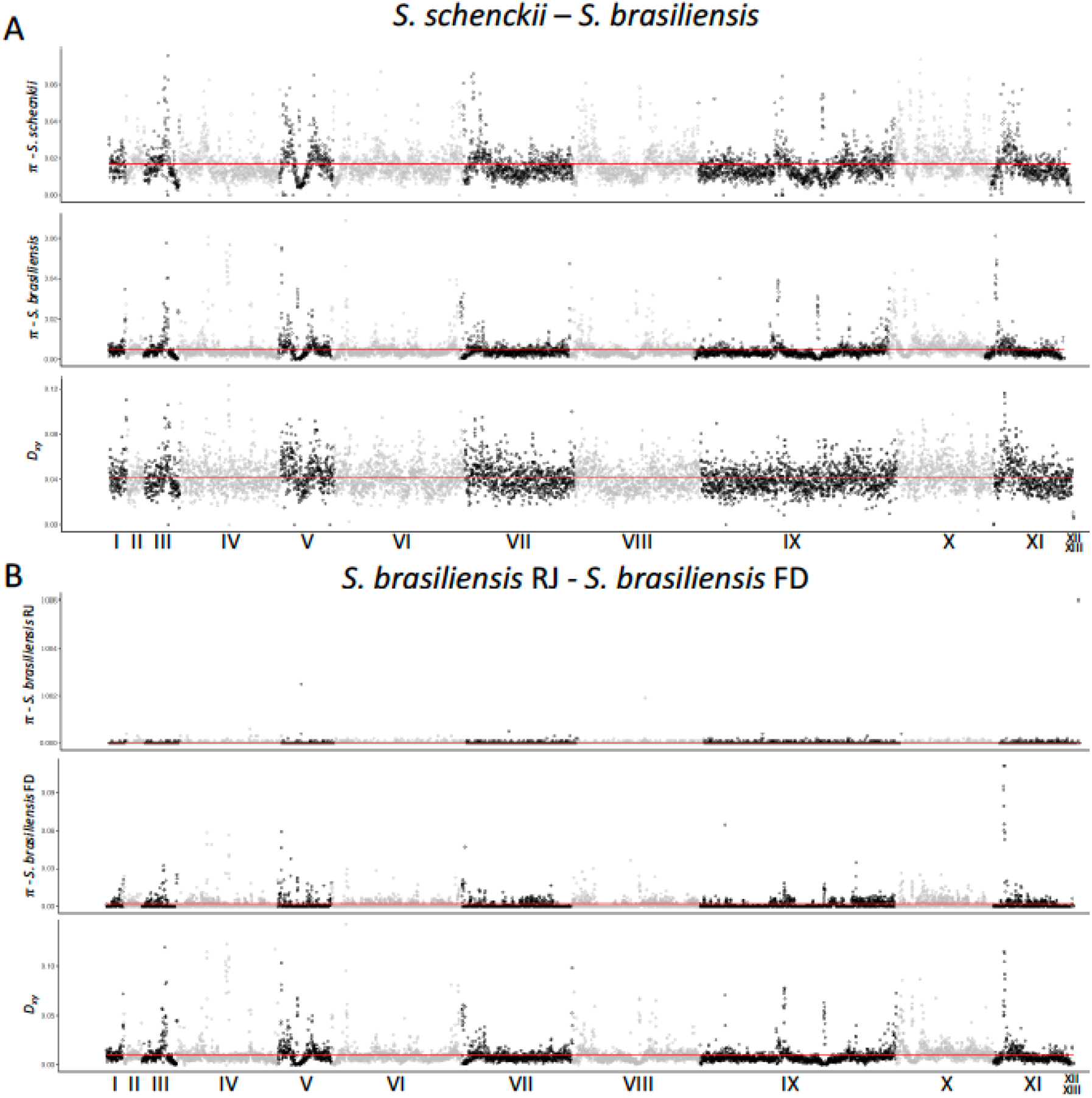
Population genetics plots along the *S. brasiliensis* 5110 reference genome. We have calculated the mean nucleotide diversity (π) within species or populations and absolute differentiation (*D*_*xy*_) between *S. brasiliensis* and *S. schenckii*, and between the *S. brasiliensis* SbFD and SbRJ (red line) and plotted along each scaffold of the *S. brasiliensis* 5110 genome using a 5kb window.

Our WGS phylogenetic tree also allows us to assess the partition of the genetic variation within *S. schenckii*. We found support for the existence of two clades, one harboring two Colombian isolates (SsEM7 and SsMS1) and a cat-derived isolate from Brasília (A003) and a clade composed by one strain from the USA (1099-18) and an additional strain from Puerto Rico (ATCC 58251). We estimate that divergence of these two groups occurred between 4.42 and 5.82 MYA. These results suggest that South American and North American populations of *S. schenckii* are genetically differentiated and call for the question of whether there has been speciation associated with geographical distribution within *S. schenckii*.

Next, we studied the relationships between the *S. brasiliensis* isolates from Brasília and Rio de Janeiro. The five Rio de Janeiro isolates (5110, s15677, s28606, s34180 and s48605) form a monophyletic group that does not include the two Brasília isolates, A001 and A005, which in turn are clustered in their own group. We also compared genomic similarities between the two *S. brasiliensis* cluster variants by aligning one individual from each cluster to the 5110 reference strains using the minimap2 tool. By aligning the *S. brasiliensis* strain s15677 to the *S. brasiliensis* 5110 reference strain (both belonging to the Rio de Janeiro population), we observed that the genomes are 99.81% similar. By aligning the *S. brasiliensis* A001 strain to the same reference, the genomic similarity dropped to 96.31% suggesting that those two groups are genetically unrelated (Supplementary Figure 1).

We calculated the approximate divergence time between *S. brasiliensis* isolates from Brasília and Southeastern Brazil. The TMRCA of the *S. brasiliensis* species was estimated to be 2.24 - 3.09 MYA, the TMRCA for the strains from Rio de Janeiro is 1.94 - 2.7 MYA, and the TMRCA for the strains from Brasília was 0.52-0.93 MYA (Figure 3). The *S. brasiliensis* isolates A001 and A005 are clustered in a single branch and are phylogenetically different from those isolated in Rio de Janeiro (5110, s15677, s28606, s34180 and s48605 – Figure 2). This result suggests that those infections were likely to be locally acquired in Brasília, not in Rio de Janeiro. These results suggest that the two populations are genetically distinct and that the infections observed in Brasília are not the result of a small group of Rio de Janeiro isolates that migrated. Since the infections caused by *S. brasiliesis* in Brasília happened autochthonously, the causal agent of these infections is genomically differentiated from the causal agent of the Rio de Janeiro infections.

We evaluate a second prediction of the hypothesis that Rio de Janeiro isolates caused the Brasília outbreak. The source population of the outbreak should have a larger genetic diversity than the outbreak population because the former has not undergone a bottleneck. We measured the magnitude of genetic variation in each *Sporothrix* group using pairwise calculations of heterozygosity. *S. schenckii* has the highest nucleotide diversity (π=1.67%), which is in line with the measurements of other fungal species with complex population composition and worldwide distribution [41,42]. The genetic variation within *S. brasiliensis* has much lower nucleotide diversity (π=0.52%) than that of its sister species, probably due to its endemicity to South America. We next calculated the nucleotide diversity within the two Brazilian *S. brasiliensis* groups. The Brasília group has a nucleotide diversity on the same order of magnitude as *S. brasiliensis* as a whole (π=0.20%). On the other hand, the Rio de Janeiro population has an extremely low genetic diversity (π=9.78 ×10^−4^%), suggesting an extreme bottleneck. In agreement with our inference from the phylogenomic tree, these results indicate that the Brasília population is extremely unlikely to be derived from the Rio de Janeiro one.

Our phylogenetic tree suggested genetic differentiation between *S. brasiliensis* isolates from Rio de Janeiro and Brasília. We calculated the pairwise absolute genetic distance (*D*_*xy*_) between *S. schenckii* and *S. brasiliensis*, and within *S. brasiliensis*. The *D*_*xy*_ between those two species was 0.042 while between the Brasília and Rio de Janeiro populations it was 0.010. Figure 3 shows the mean pairwise differences per site within populations (nucleotide diversity or π) and mean pairwise differences per site between populations *D*_*xy*_ along the 13 scaffolds of the *Sporothrix* genome. The genome differentiation between *S. schenckii* and *S. brasiliensis* is extensive and widespread along the whole genome. We observed a strikingly different pattern when we compare the two populations of *S. brasiliensis*. As expected, Rio de Janeiro *S. brasiliensis* has little genetic variation along the whole genome (Figure 3A). The *D*_*xy*_ along most of the genome is low suggesting low differentiation between Rio de Janeiro and Brasília despite almost 1 MY of divergence (See above, Figure 2). Nonetheless, scaffolds I, III, V, IX and XI, show peaks of high differentiation. Notably, the *D*_*xy*_ peaks in scaffolds I, III, and XI are highly differentiated between Rio de Janeiro and Brasília *S. brasiliensis* and between *S. brasiliensis* (as a whole) and *S. schenckii*. These results suggest that the differentiation between Brazilian populations of *S. brasiliensis* is modest and localized and not as pronounced as between *S. schenckii* and *S. brasiliensis*. This difference might simply stem from the difference in divergence time between the *Sporothrix* species and the *S. brasiliensis* populations.

## Discussion

Sporotrichosis due to *S. brasiliensis* has become one of the most important fungal diseases in Brazil. The disease has gone from a localized outbreak in Rio de Janeiro state to a country-wide epidemic during the past 20 years and has affected thousands of humans and animals. In Brasília, located in Midwestern Brazil and roughly 900 km from Rio de Janeiro, we observed indications that suggest cat/dog-transmitted sporotrichosis might occur. Moreover, both cats and dogs are affected by pathogenic *Sporothrix* spp. In Rio de Janeiro, both fungal species have been isolated from dogs [18], but *S. brasiliensis* has been identified in almost all cats tested so far [16]. The etiologic agent of this disease was recently found in clinical and environmental samples in Argentina as well in veterinary samples in Paraguay [19,20].

In this study, we dissect the support for two different hypotheses for the expansion of this disease throughout South America, namely that *S. brasiliensis* recently (< 20 years) spread from Rio de Janeiro to other areas in Brazil and other countries via human, animal hosts or fomites; or that different groups of *S. brasiliensis* occur in different locations of the South American continent. We focused on a recent sporotrichosis case series in Brasília. If the former hypothesis is correct, one would expect that the Rio de Janeiro population would have a larger genetic variation than the Brasília population. Additionally, one would expect the Rio de Janeiro population to encompass the genetic variants observed in Brasília. Neither of these were observed in our data. The nucleotide diversity (π) in Rio de Janeiro is 200 times lower than in Brasília. Additionally, the Rio de Janeiro and Brasília populations are reciprocally monophyletic which contradicts the idea that Brasília strains are a subset of the Rio de Janeiro group. Moreover, the split time between Brasília and Rio de Janeiro occurred between 0.52-0.93 MYA, predating the human colonization of the Americas and thus precluding the possibility that *S. brasiliensis* migrated associated with humans.

The observation that the surge on sporotrichosis in Brasília is not caused by recent migration from Rio de Janeiro poses two additional questions. First, other reasons, besides migration must be responsible for the recent uptick of cases. One possibility is that deforestation and human expansion into previously uninhabited areas has put humans (and their pets) in contact with fungi that otherwise had never infected humans. A second possibility is that in the last 20 years, a previously tamed lineage of *S. brasiliensis* became more virulent. Both novel mutation and introgression from a virulent lineage could be responsible. Finally, it is possible that the improvement of epidemiology and medical observations has led to the identification and recording of sporotrichosis due to *S. brasiliensis*.

Second, it remains unclear why the number of cases in Brasilia is lower than that in Rio de Janeiro, one of the most hyperendemic areas in the world. One potential explanation for the higher incidence of sporotrichosis in Rio de Janeiro than Brasília is higher population density and less favorable sanitation conditions in the former area [17]. Our study shows that cats can acquire sporotrichosis caused by both *S. schenckii* and *S. brasiliensis*, which indicates that feral and stray cats can be reservoirs of the disease. Another possibility is that Brasilia and Rio de Janeiro differ in their number of outdoor cats. Studies have shown a high prevalence of parasitosis in cats from both locations (Brasilia: [43], RJ:[44]) but the number of felines and their relative health conditions remain unknown.

Our work suggests the existence of at least two groups of *S. brasiliensis* in South America. The increase in sporotrichosis incidence, along with the increments on the disease burden of other mycoses across the world, and the recent outbreak of *Cryptococcus gattii* - originated in the Amazon region - in the Pacific Northwest [45] are sobering indications of the importance of efforts to detect and mitigate or prevent the spread of fungal emerging diseases, both in South America and elsewhere.

## Data Availability

The data presented in this manuscript is either available on public databases, such as GenBank, or is available upon request to the authors.

## Acknowledgments

The authors would like to thank to Jéssica S. Boechat and Manoel M. E. Oliveira (Fiocruz) for the technical support. A.M.N was funded by FAP-DF awards 0193.001048/2015-0193.001561/2017 and the CNPq grant 437484/2018-1. B.M.B. was supported by NIH/NIAID award R21AI28536. D.R.M. was supported by NIH/NIGMS award R01GM121750. M.M.T was supported by CNPq/UNIVERSAL award 43460/2018-2. M.S.S.F was supported by FAP-DF/PRONEX award 193.001.533/2016. S.A.P. was supported by FAPERJ, grant number E-26/202.737/2019.

---

**Supplementary Figure 1.**
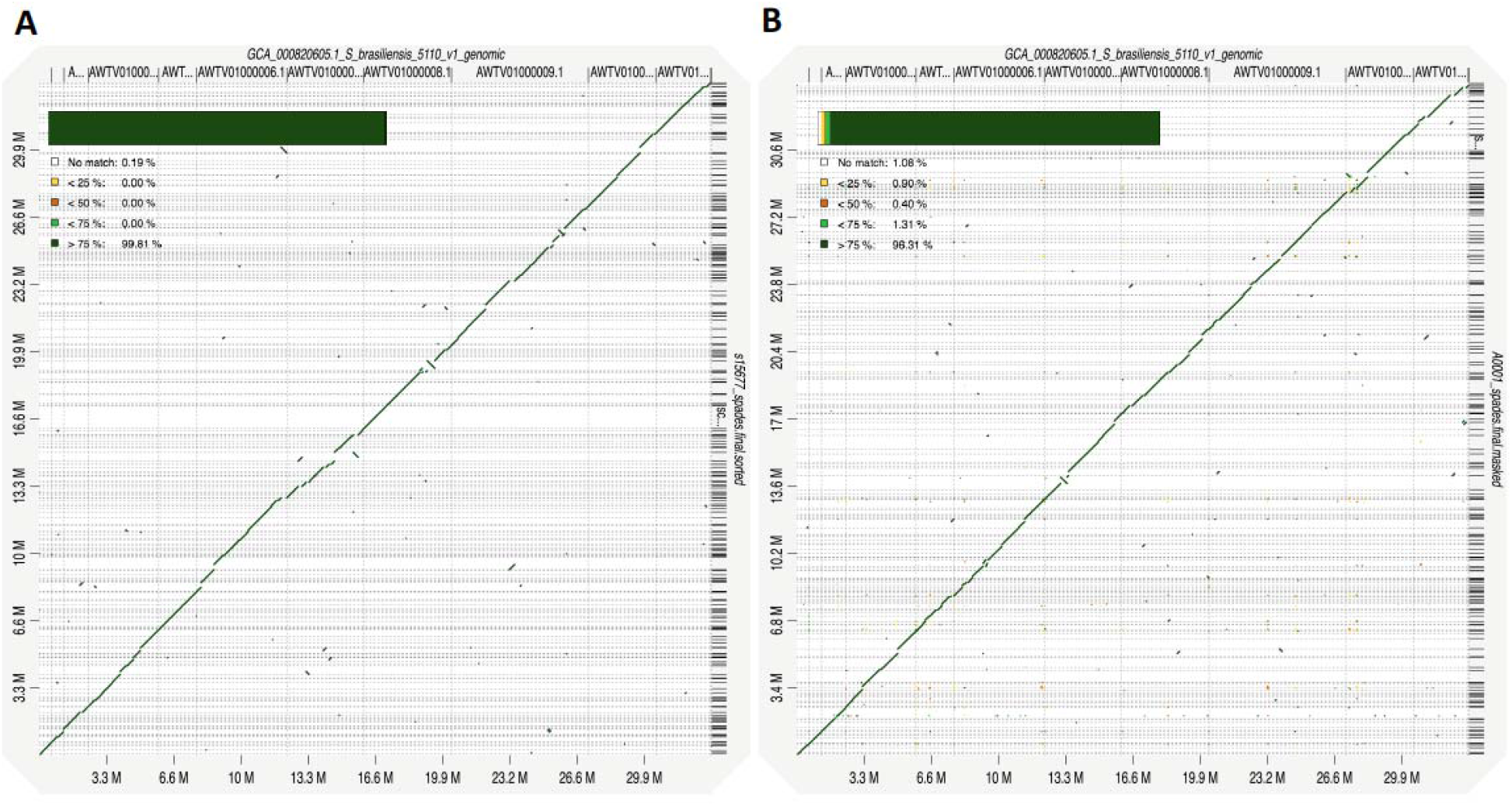
Dot plots of whole genome alignments between different *S. brasiliensis* genotypes. The *S. brasiliensis* strains s15677 (A - RJ population) and A001 (B – FD population) were aligned to the *S. brasiliensis* 5110 reference strain and the similarity values are displayed for each scenario.

